# Armed Violent Conflict and Healthcare-Seeking Behavior for Maternal and Child Health in Sub-Saharan Africa: A Systematic Review

**DOI:** 10.1101/2024.03.11.24304112

**Authors:** Gbadebo Collins Adeyanju, Pia Marie Elisabeth Schrage, Rabiu Ibrahim Jalo, Liliana Abreu, Max Schaub

**Author notes:** **Corresponding Author:** Gbadebo Collins Adeyanju Center for Empirical Research in Economics and Behavioral Science (CEREB) University of Erfurt, Erfurt, Germany.

## Abstract

**Background:** Over 630 million women and children worldwide face displacement due to conflict or resided dangerously close to conflicts zones. While the adverse effects of physical destruction on healthcare delivery are relatively well understood, the effects on healthcare-seeking behavior remain underexplored, especially in sub-Saharan Africa. This study aims at the interconnections and knowledge gaps between exposure to armed violent conflicts and healthcare-seeking behaviors for maternal and child health in Sub-Saharan Africa.

**Methods:** Five key electronic databases (PubMed, Scopus, Web of Science, PsycNET, and African Journals Online) were **s**earched for peer-reviewed publications between 2000 and 2022. The review was designed according to PRISMA-P statement and the protocol was registered with PROSPERO database. The methodological quality and risks of bias were appraised using GRADE. A data extraction instrument was modelled along the Cochrane Handbook for Systematic Reviews and the Centre for Reviews and Dissemination of Systematic Reviews.

**Result:** The search results yielded 1,148 publications. Only twenty-one studies met the eligibility criteria, reporting healthcare-seeking behaviors for maternal and child health. Among the twenty-one studies, seventeen (81.0%) reported behaviors for maternal health such as antenatal care, skilled birth attendance, postnatal care services, and family planning. Similarly, nine studies (42.8%) observed behaviors for child health such as vaccination uptake, case management for pneumonia, diarrhea, malnutrition, and cough. While conflict exposure is generally associated with less favorable healthcare-seeking behavior, in some of the studies, healthcare outcomes improved. Marital status, male partner’s attitude, education, income and poverty levels were associated with healthcare-seeking behavior.

**Conclusion:** There is need for multifaceted interventions to mitigate the repercussions of armed violent conflicts on healthcare-seeking behavior, given its mixed effects on child and maternal healthcare utilization. While armed violent conflict disproportionately affects child compared to maternal health, it is noteworthy that, exposure to such conflicts may unintentionally also lead to positive outcomes.

**Prospero registration number:** CRD42023484004.

## INTRODUCTION

The health status of populations, especially women and children, tends deteriorate in countries affected by armed conflicts. Exposure to armed conflicts often leads to an overwhelming loss of human life, disruptions of health service delivery, infrastructure breakdown and widespread displacement [1]. The International Committee of the Red Cross (ICRC) classifies armed conflicts as non-international conflicts involving governmental and non-government forces, these include insurgency, organized crimes and banditry [2]. Meanwhile, the Uppsala Conflict Data Program (UCDP) sees armed conflict as a contested incompatibility related to government and/or territory, wherein the use of armed force between the military forces of two parties, with at least one being a government of a state, has resulted in at least 25 battle-related deaths each year [3].

A substantial portion of the world’s women and children resides in countries suffering from armed conflicts [4]. Estimates indicate that, in 2017, approximately 630 million women and children, accounting for 10% of women and 16% of children worldwide, either faced displacement due to conflict or resided dangerously close to conflicts zones [4,5,6,7]. The problem is particularly pronounced in sub-Saharan Africa (SSA), where armed conflicts have been more prevalent than in other regions. For example, nearly 70% of all countries in SSA have witnessed armed conflicts since 1980 [4,8].

It is widely recognized that the impact of wars extends far beyond the cease fire [9–11]. However, the underlying reasons for this phenomenon are yet to be fully understood. Some adverse long-term effects can be directly attributed to the conflict itself: unexploded ordnance continues to pose a threat to lives and livelihoods even decades after a war [12], and injuries sustained during conflicts shorten lives and compromises the health of those affected [13]. Wars cause health consequences beyond the physical injuries resulting from violence: the destruction of sanitary and energy infrastructure can lead to the spread of water-borne and respiratory diseases [14]. Reduced functioning or restricted access to health facilities may reduce vector control and contribute to the spread of mosquito-transmitted diseases such as Malaria or Dengue fever [15,16]. Refugees and internally displaced individuals may inadvertently introduce diseases to other areas [17,18]. The destruction of hospitals and clinics, combined with the departure of health workers seeking safety for themselves and their families, further exacerbates healthcare challenges in conflict zones [19–23].

Beyond tangible impacts, armed violent conflicts can have intangible effects, including the normalization of violence, shifts in social norms, and the breakdown of the social order. These elements are often accompanied by increased levels of sexual violence and voluntary unprotected sex, leading to the spread of sexually transmitted diseases [24]. The trauma resulting from violence, even if not visibly apparent, is associated with both persistent mental health issues and associated physical ailments that often afflict victims throughout their lives [11]. Moreover, exposure to violence may lead to fear, mistrust, and psychological trauma, creating reluctance among individuals to seek out health-services [25].

This study aims to generate new insights into the complex interplay between health decision-making or health-seeking behavior and exposure to armed violence during and after conflicts. It seeks to generate knowledge that can be used in health promotion initiatives in conflict and post-conflict societies, especially those that aim at reducing child and maternal mortality and morbidity. The specific objectives of this study are to assess the existing body of knowledge on healthcare-seeking behaviors in armed violent conflict settings, and to explore and describe the various factors influencing healthcare-seeking behaviors in regions of Sub-Saharan Africa affected by armed violent conflicts.

## METHODS

### Systematic review registration and reporting

The review protocol was registered with the PROSPERO database (Reg. no.: CRD42023484004) and designed in accordance with the PRISMA 2020 Statement [26].

### Data Sources and Search Strategy

An interdisciplinary review team with expertise in global health, public health, public policy, epidemiology, political science and conflict studies, social science and evidence synthesis was constituted. Guided by the Population, Intervention, Comparison and Outcomes (PICO) model for systematic review [27,28] (see Table 1), the team developed a broad research question: *“How does exposure to armed violent conflicts affect healthcare-seeking behaviors for maternal and child health in sub-Saharan Africa?”*.

**Table 1:** The PICO framework for systematic review.

| **PICO** | **Measure** | **Keywords** |
| --- | --- | --- |
| **Population** | Parents or caregivers of under-fives, pregnant women, and women in SSA | Children, child, maternal, prenatal, antenatal, pregnancy, all countries in SSA. |
| **Intervention or exposure** | Experience of (exposure to) armed violent conflicts | War, armed conflict, conflict experiences, conflict exposure, organized violence, armed violence, fighting, insurgency, war trauma, traumatic experiences, civil conflict, conflict-affected populations, war-affected communities |
| **Comparator** | Not Applicable | Not Applicable |
| **Outcome** | Healthcare-seeking behavior | Healthcare-seeking behaviour; healthcare-seeking behavior; health service utilization; health service utilization; healthcare utilization; healthcare utilisation; healthcare access; access to healthcare; health behavior; health behaviour; prenatal care; postnatal care; healthcare decision-making, healthcare choices, healthcare barriers, delayed care; refused healthcare; healthcare; seeking; behaviour; behavior; health; service; utilization; utilisation; use of maternal health services, use; maternal; services; healthcare access; access to healthcare; utilization of healthcare services; utilisation of healthcare services |

A systematic search was conducted on four general and one key electronic database. The general databases were: MEDLINE via PubMed Central, Scopus, Web of Science and PsycNET. Because of its rich repository of Africa-based journal publications, we also included the African Journals Online (AJOL). Suitable MeSH and thesaurus terms were adapted with key terms in each database searched. Three strings of multiple combinations of search terms and relevant Boolean operators to the study PICO framework were developed as seen in Table 2, such as Population (parents and caregivers of under-5, children under-5, pregnant women, and mothers in SSA), Intervention (experience of or exposure to armed violent conflicts), Comparison (none) and Outcome (healthcare-seeking behavior: e.g., maternal and child health). Two independent reviewers conducted the initial screening of the titles, abstracts, and keywords of relevant studies, prior to the review of the full text involving all authors.

**Table 2:** Search strategy: #1, #2, and #3.

|  |  |  |
| --- | --- | --- |
| **Search string #1** | **Search string #2** | **Search string #3** |
| "armed conflict"<br>OR "armed<br>violence" OR<br>"insurgency" OR<br>"conflict<br>experiences" OR<br>"conflict<br>exposure" OR<br>"war trauma" OR<br>"traumatic<br>experiences" OR<br>"civil conflict" OR<br>"conflict-affected<br>populations" OR<br>"war-affected<br>communities"<br>AND | "maternal health" OR<br>"child health" OR<br>"reproductive health<br>services" OR "childhood<br>health" OR "maternal<br>health services" OR<br>"healthcare-seeking<br>behavior" OR "health<br>service utilization" OR<br>"healthcare" OR<br>"seeking" OR "behaviour"<br>OR "behavior" OR<br>"health" OR "service" OR<br>"utilization" OR "use of<br>maternal health services"<br>OR "use" OR "maternal"<br>OR "services" OR<br>"healthcare access" OR<br>"access to healthcare" OR<br>"healthcare utilization"<br>OR "utilization of<br>healthcare services" OR<br>"healthcare decision-making" OR "healthcare<br>choices" OR "healthcare<br>barriers" AND | "Africa"[MeSH] OR Africa*[tw] OR Algeria[tw] OR<br>Angola[tw] OR Benin[tw] OR Botswana[tw] OR<br>"Burkina Faso"[tw] OR Burundi[tw] OR Cameroon[tw]<br>OR "Cape Verde"[tw] OR "Central African Republic"[tw]<br>OR Chad[tw] OR Comoros[tw] OR Congo[tw] OR<br>"Democratic Republic of Congo"[tw] OR Djibouti[tw]<br>OR "Equatorial Guinea"[tw] OR Eritrea[tw] OR<br>Ethiopia[tw] OR Gabon[tw] OR Gambia[tw] OR<br>Ghana[tw] OR Guinea[tw] OR "Guinea Bissau"[tw] OR<br>"Ivory Coast"[tw] OR "Cote d'Ivoire"[tw] OR Kenya[tw]<br>OR Lesotho[tw] OR Liberia[tw] OR Madagascar[tw] OR<br>Malawi[tw] OR Mali[tw] OR Mauritania[tw] OR<br>Mauritius[tw] OR Mayotte[tw] OR Mozambique[tw] OR<br>Mocambique[tw] OR Namibia[tw] OR Niger[tw] OR<br>Nigeria[tw] OR Principe[tw] OR Reunion[tw] OR<br>Rwanda[tw] OR "Sao Tome"[tw] OR Senegal[tw] OR<br>Seychelles[tw] OR "Sierra Leone"[tw] OR Somalia[tw]<br>OR "South Africa"[tw] OR "St Helena"[tw] OR<br>Sudan[tw] OR Swaziland[tw] OR Tanzania[tw] OR<br>Togo[tw] OR Uganda[tw] OR "Western Sahara"[tw] OR<br>Zaire[tw] OR Zambia[tw] OR Zimbabwe[tw] OR "Central<br>Africa"[tw] OR "Central African"[tw] OR "West<br>Africa"[tw] OR "West African"[tw] OR "Western<br>Africa"[tw] OR "Western African"[tw] OR "East<br>Africa"[tw] OR "East African"[tw] OR "Eastern<br>Africa"[tw] OR "Eastern African"[tw] OR "South<br>African"[tw] OR "Southern Africa"[tw] OR "Southern<br>African"[tw] OR "sub Saharan Africa"[tw] OR "sub<br>Saharan African"[tw] OR "subSaharan Africa"[tw] OR<br>"subSaharan African"[tw] OR Africa South of the<br>Sahara[MH:EXP] NOT ("guinea pig"[tw] OR "guinea<br>pigs"[tw] OR "aspergillus niger"[tw]) |

### Study selection process

The search results were imported into the Zotero reference management software to compile and filter relevant articles that meet the eligibility criteria and to exclude duplicates [29]. In the initial stage, five authors (GCA, LA, MS, PS, and RIJ) independently reviewed all articles using titles, keywords, and abstracts. This was done to determine their alignment with the article’s suitability to the goal of the review and based on defined inclusion and exclusion criteria. The second stage involved scanning full-text articles of all included studies to determine final eligibility. All included studies from the first and second stages were proportionally distributed and assessed strictly against the inclusion and exclusion criteria described in Table 3. Disagreements or non-aligned decisions were resolved through weekly workshops with all authors. The PRISMA flow chart for systematic review was used to document the study selection process (see Figure 1).

**Figure 1:**
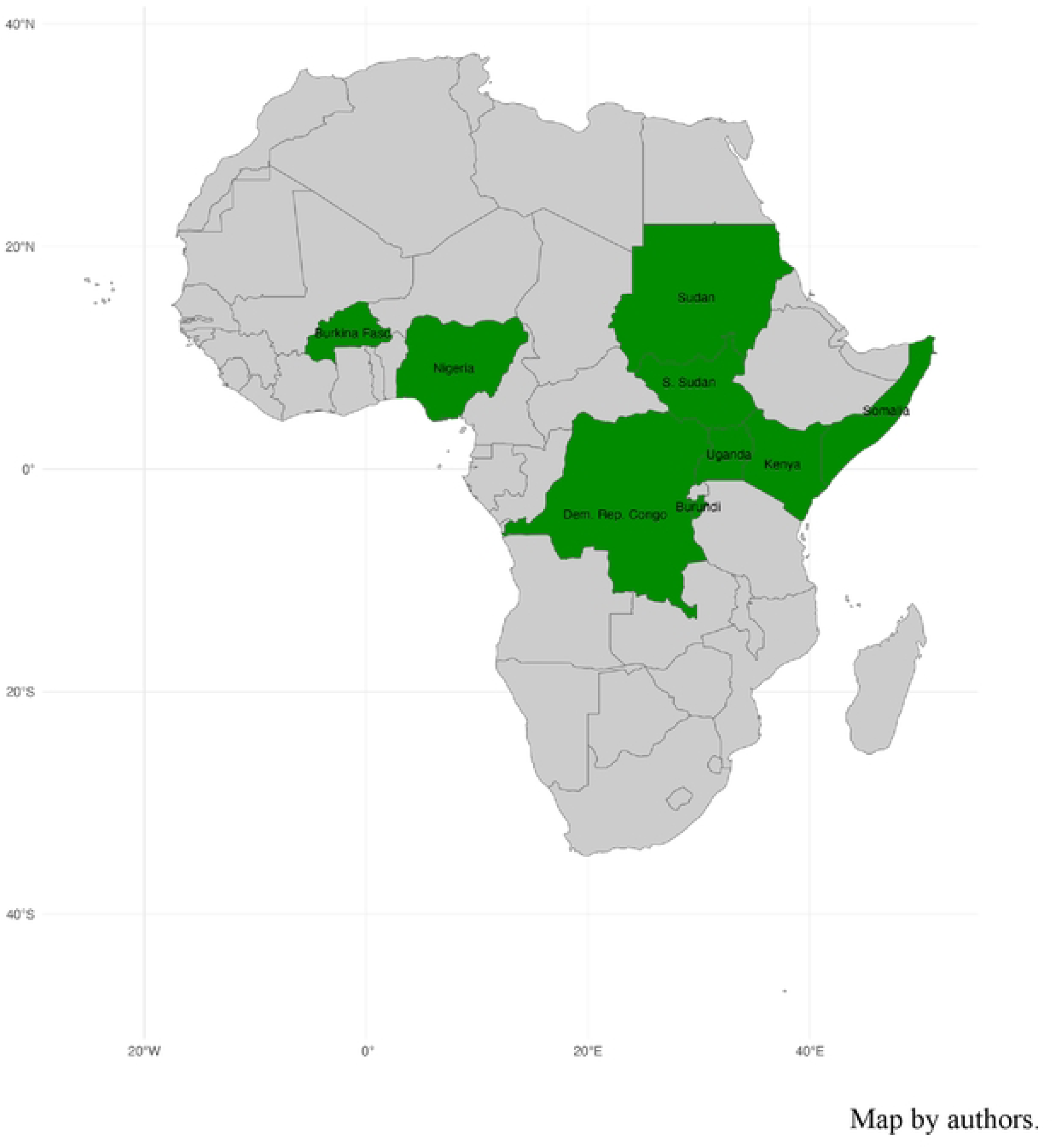
Eligible studies by country across SSA.

**Table 3:** Inclusion and exclusion criteria.

| Inclusion | Exclusion |
| --- | --- |
| a. English language only.<br>b. Only publications between 2000 - 2022<br>c. All countries within the sub-Saharan Africa region only, as defined by United Nations development Program [28].<br>d. All studies that assess experience or exposure to armed violent conflicts and its consequent impact on health outcomes.<br>e. All studies with outcomes significantly impacting behavior and decision-making, leading to changes in health.<br>f. Only studies which assess maternal and child health. | a. Published research articles in non-peer-reviewed journals.<br>b. Studies focused on gender-based or domestic violence.<br>c. Studies whose primary or secondary outcomes are not healthcare-seeking behavior or health decision-making.<br>d. Studies on the cost of healthcare in armed conflict settings<br>e. Studies that report findings only on health behavior or health changes not influenced by armed violent conflict |

### Study Eligibility

Studies were included in the review according to the PICO criteria. To avoid distractions associated with non-relevant studies, a predefined inclusion and exclusion criteria was developed (see Table 3).

### Outcome of interest

Since studies on healthcare-seeking behaviors include a diverse demographic spectrum, ranging from infants to the elderly, this study targeted parents/legal guardian (caregivers) of children under the age of five years, pregnant women, and mothers in SSA. Healthcare-seeking behavior has been defined as any action taken by individuals who perceive themselves to have a health problem or are ill, for the purpose of finding an appropriate remedy [31]. Given the comprehensive nature of healthcare-seeking behavior, this review focuses specifically on the utilization of health services in armed conflict settings.

The impacts of exposure to or experience of armed violence conflict on health-seeking behavior in SSA were classified into two broad categories. Maternal health variables include antenatal care attendance, maternal vaccination, skilled attendance at birth, and postnatal care services (e.g., utilization of family planning services). Child health variables include the uptake of childhood vaccination from routine immunization systems, and healthcare-seeking behaviors for common health problems among children under the age of five years, such as fever and diarrhea disease.

### Quality assessment of studies and risk of bias

The quality of evidence in each study in the review was assessed using the Grades of Recommendation, Assessment, Development, and Evaluation Working Group [32]. The review sided with the World Health Organization (WHO) and Cochrane Collaboration on the principles of the GRADE system for evaluating the quality of evidence for outcomes reported in systematic reviews [33]. The authors independently assessed each relevant study using the criteria from the Cochrane Collaboration and the Centre for Reviews and Dissemination (CRD) to evaluate the risk of bias and quality of evidence [34].

### Data Extraction

A data extraction instrument was developed using Microsoft Excel, based on the Cochrane handbook for Systematic Reviews and the CRD’s Guidance for Undertaking Reviews in Health Care [35,36]. Two algorithms were formulated. The first addressed the characteristics of study findings. It comprised key indicators such as author last name, publication year, title of publication, country of study, study design, primary and secondary outcomes, nature of armed violence conflict, study setting, gender of the targeted group, and factors associated with health-seeking behavior that were reported in the studies, especially those related to maternal and child health. The second algorithm was used to assess the quality of evidence and risks of potential biases in each study, such as study design, selection, detection, reporting, attrition, and publication bias. In addition, the assessment considered evidence related to impression, inconsistency, and indirectness in the studies. The factors influencing health-seeking behavior identified in the study due to exposure to armed violent conflicts were systematically categorized into themes. Two authors (PS and RIJ) independently extracted data from the final included studies, and populated the two designed matrices, which were then peer-reviewed in a workshop among all authors to confirm the collected data and resolve disagreements or misinterpretations.

### Study Analysis

Significant evidence relevant to the goal of the review was extracted, appraised and reported in a systematic fashion using the PRISMA-P checklist [37,38]. The extracted data were synthesized to answer the research question.

## RESULTS

Figure 1 visualizes the country-contexts of the final set of studies that met all the review criteria. All the included nine countries (Nigeria, Burkina Faso, DRC, Sudan, South Sudan, Somalia, Uganda, Kenya, and Burundi) have experienced prolonged armed violent conflicts over several decades. In Somalia, since the formation of the Al Qaeda-affiliated Al-Shabaab group, at least 1,000 deaths have been reported every year [39]. Presently, widespread conflict has resulted in the deterioration of Somalia’s public health system with over 2.6 million internally displaced persons due to the conflict and drought [39,40]. In eastern DRC, nearly two decades of armed conflict have significantly compromised the health system, contributing to an estimated 3.9 million excess deaths [41,42]. Uganda is currently recovering from over 20 years of armed conflict orchestrated by the Lord’s Resistance Army, which resulted in the disruption of health services and massive population displacement estimated at 2 million [43,44]. Both Burundi and Uganda are recovering from atrocious civil wars that each claimed tens of thousands of lives and caused millions of people to be displaced [45]. In West African countries like Nigeria and Burkina Faso, constant conflicts due to religion, ethnicity, and political views have posed significant challenges [43]. Currently, Boko Haram operating in Nigeria is recognized as one of the most extreme terrorist groups in SSA. Since 2009, the Boko Haram insurgency has killed more than 20,000 people and displaced more than 2 million [46].

Nigeria, Somalia, the DRC, Burkina Faso, Sudan, South Sudan, and Cameroon have often experienced very high-intensity armed conflicts, while conflicts tend to be of lower intensity in countries such as Kenya and Uganda. These conflicts involved non-state actors, terrorists, bandits, and other armed groups. Overall, the intricate humanitarian emergencies caused by armed violent conflicts severely impact the already fragile national and sub-national health systems in many of these countries through the destruction of health facilities and flight of trained health workers [47,48].

### Search results

The search across five databases [PubMed, Scopus, Web of Science, PsycNET, and African Journals Online (AJOL)] resulted in 1,148 articles (PubMed = 596, Scopus = 218, Web of Science = 137, PsycNET = 159 and AJOL = 38), with 77 articles removed as duplicates. Therefore, 1,071 articles were then screened based on title and abstract for eligibility. A total of 1,005 articles were excluded during this screening phase for various reasons: articles not in English (n=3), not peer-reviewed (n=17), health-seeking behavior not the primary outcome (n=627), not focusing on armed conflict (n=30), not studies in SSA (n=86), studies of sexual or domestic violence (n=91), studies of healthcare costs (n=10), studies on internally displaced persons or refugees (n=44), review articles (n=36), and finally, studies whose focus was not maternal and child health (n=61).

The remaining 66 articles underwent a full text screening. After the screening, 45 articles were excluded based on: duplicates (n=2), not peer-reviewed article (n=1), review articles (n=8), outcome not defined (n=2), health-seeking behavior not the primary outcome (n=20), not focused on armed conflict (n=3) and maternal and child health (n=9), respectively. The final review incorporated 21 studies reporting healthcare-seeking behaviors for maternal and child health in conflict and post-conflict settings in SSA. The screening and selection process is illustrated in the PRISMA-P flow diagram of Figure 2.

**Figure 2:**
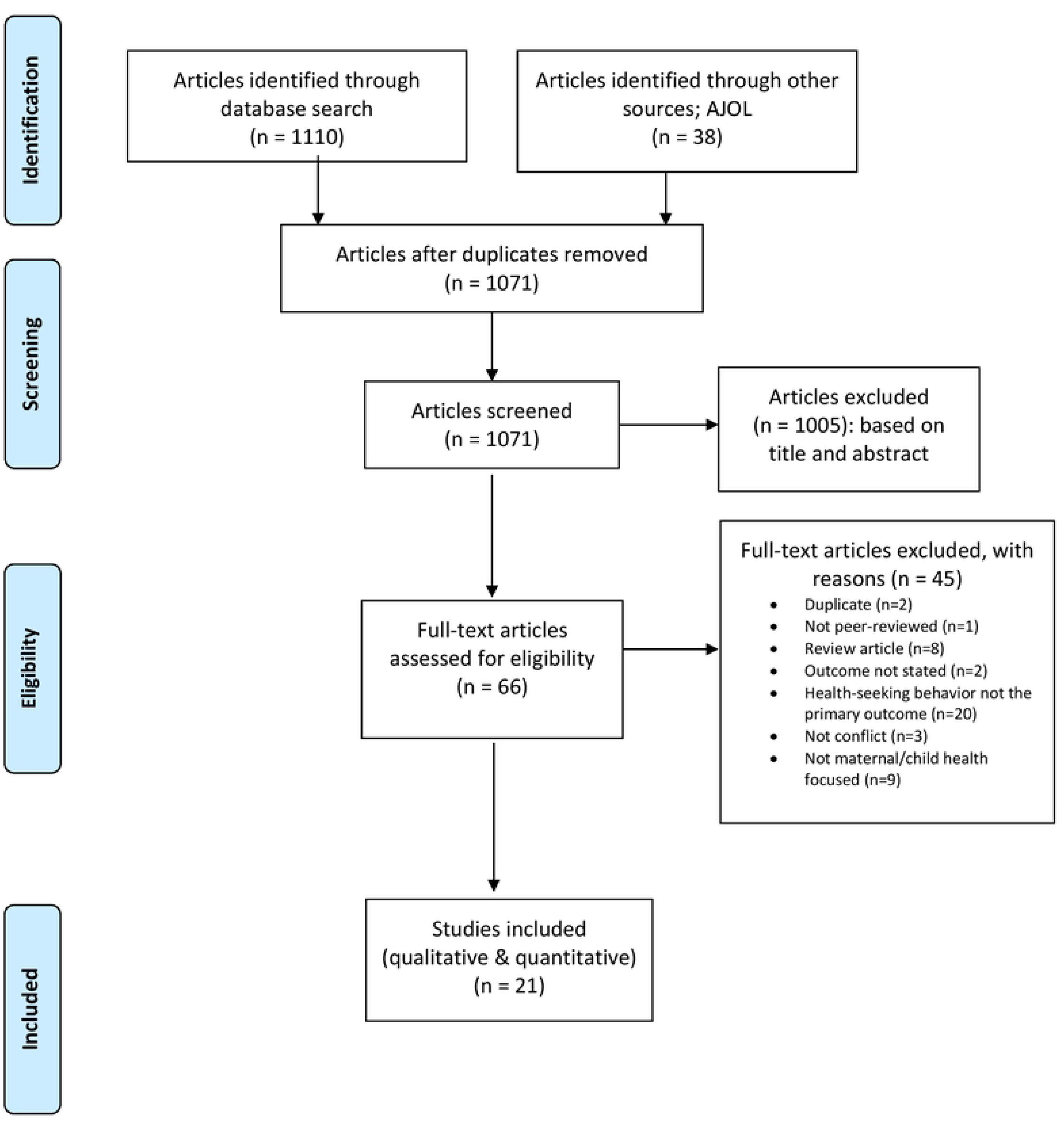
PRISMA 2009 Flow Diagram [49]

A significant proportion of the eligible studies were from Nigeria (n=8, 38.1%), Uganda (n=3, 14.2%), and Congo (n=3, 14.2%). All studies were peer-reviewed journal articles. Out of the total, 13 studies (61.8%) were descriptive in design, while three (14.3%) used a pre-post survey approach. 12 studies were on maternal health issues only (57.2%) and four (19.0%) involved children exclusively, while five studies reported outcomes for both children and women (23.8%). The majority of the studies (90.5%) were published within the last ten years (2012-2022), and none were randomized control trials. The characteristics of the included studies are presented on Table 4.

**Table 4:** Characteristics of included studies.

| **Variable** | **Frequency** | **Percentage (%)** |
| --- | --- | --- |
| **Year of study** |  |  |
| Before 2012 | 2 | 9.5 |
| 2012 and after | 19 | 90.5 |
| **Study design** |  |  |
| Cross sectional | 13 | 61.8 |
| Pre – post | 3 | 14.3 |
| Quasi-experimental | 1 | 4.8 |
| Longitudinal | 1 | 4.8 |
| Others | 3 | 14.3 |
| **Study setting** |  |  |
| Single country | 18 | 85.7 |
| Multi-country | 3 | 14.3 |
| **Type of health seeking behavior** |  |  |
| Maternal | 12 | 57.2 |
| Child | 4 | 19.0 |
| Maternal & child | 5 | 23.8 |
| **Nature of violent conflict experienced** |  |  |
| Armed conflict | 7 | 33.3 |
| Civil conflict | 5 | 23.8 |
| Insurgency | 9 | 42.9 |
| **Country of study** |  |  |
| Nigeria | 8 | 38.1 |
| Somalia | 1 | 4.8 |
| South Sudan | 1 | 4.8 |
| Uganda | 3 | 14.2 |
| Congo (DRC) | 3 | 14.2 |
| Burundi & Uganda | 2 | 9.5 |
| Burkina Faso | 1 | 4.8 |
| Sudan, Uganda & DRC | 1 | 4.8 |
| Kenya | 1 | 4.8 |
| **Study method** |  |  |
| Quantitative | 13 | 61.9 |
| Qualitative | 3 | 14.3 |
| Mixed methods | 5 | 23.8 |

As outlined in Table 5, the included studies reported different outcomes for healthcare-seeking behaviors in SSA. Of the twenty-one included studies, seventeen (81.0%) reported healthcare-seeking behaviors for maternal health services in settings of armed violent conflict. The reported healthcare-seeking behaviors were antenatal care attendance, skilled birth attendance, postnatal care services, family planning uptake/contraceptive prevalence rate, health facility birth, caesarean section rates, unmet need for family planning, and HIV/AIDs care and treatment [19,20,39,41,45,47,50–60]. Nine studies (42.9%) reported healthcare-seeking behaviors for child health. The child health seeking behaviors reported were vaccines uptake (measles, BCG, and DPT), and case management for pneumonia, diarrhea, malnutrition, fever, and cough [39,43,50,56,58,59,61–63]. Antenatal care attendance was the most common healthcare behavior observed in most of the studies (57.1%), followed by utilization of skilled birth attendants (33.3%) for maternal health, while the uptake of measles vaccination was the most common health-seeking behavior for child health.

**Table 5:**
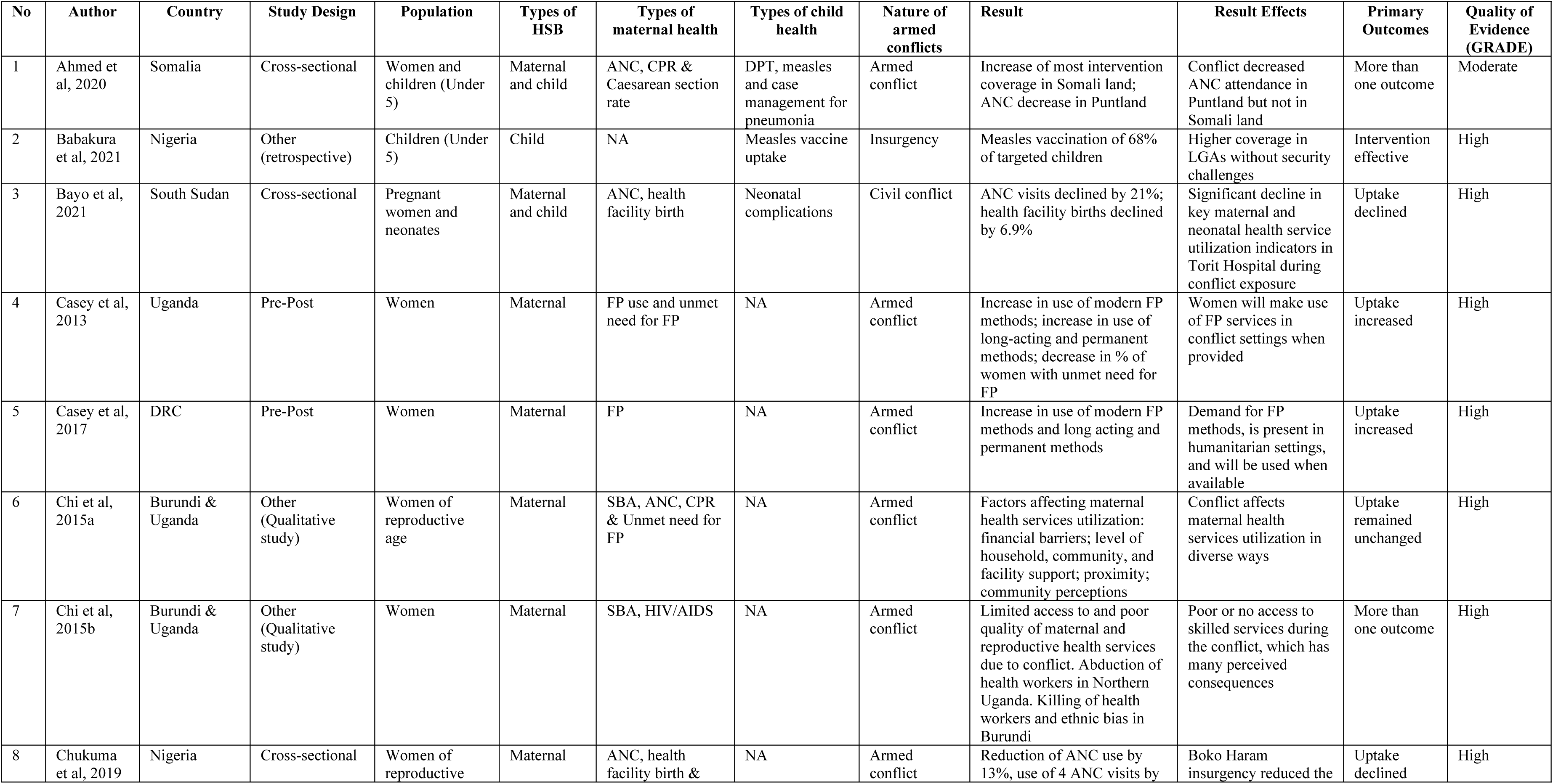
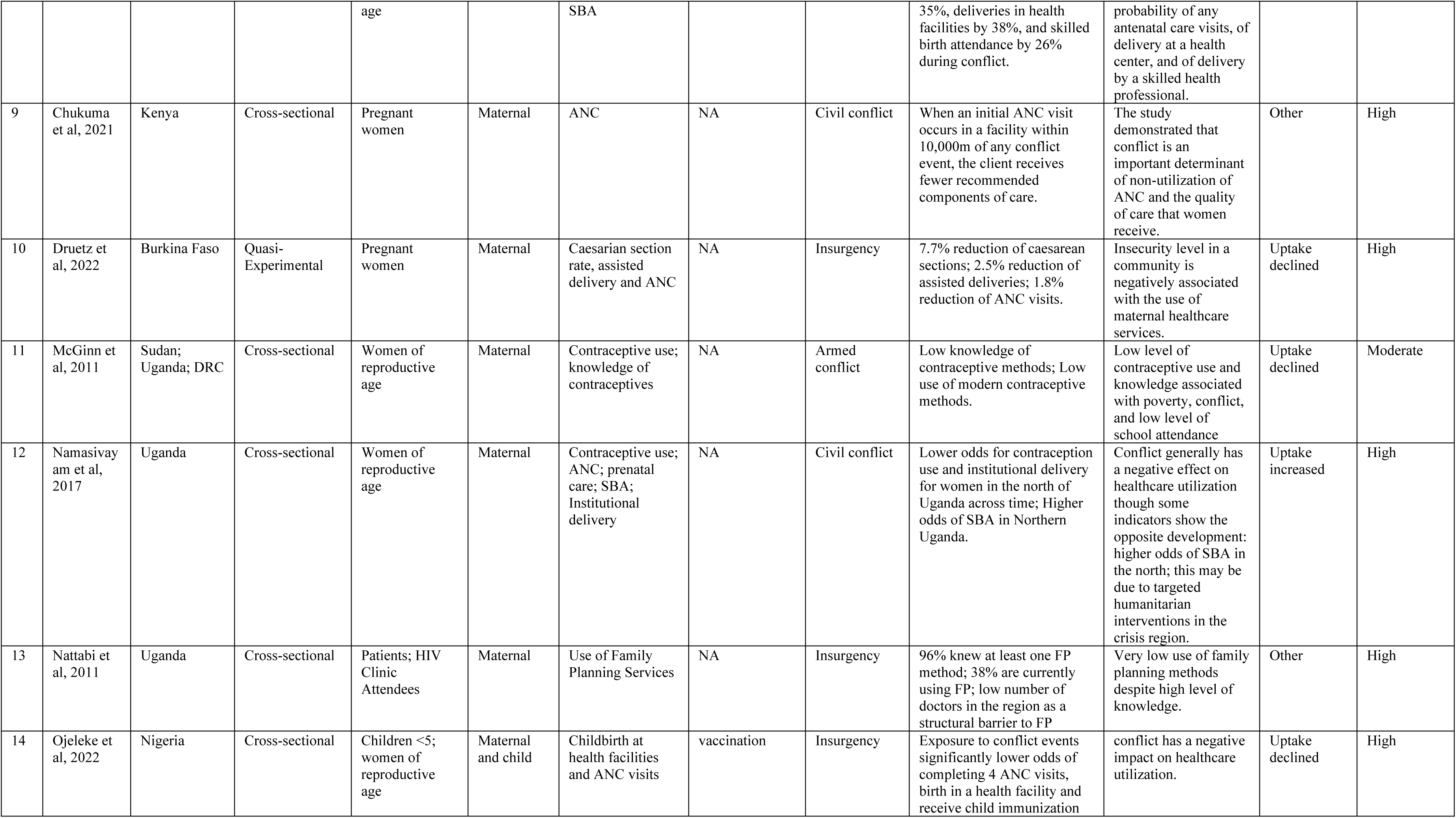
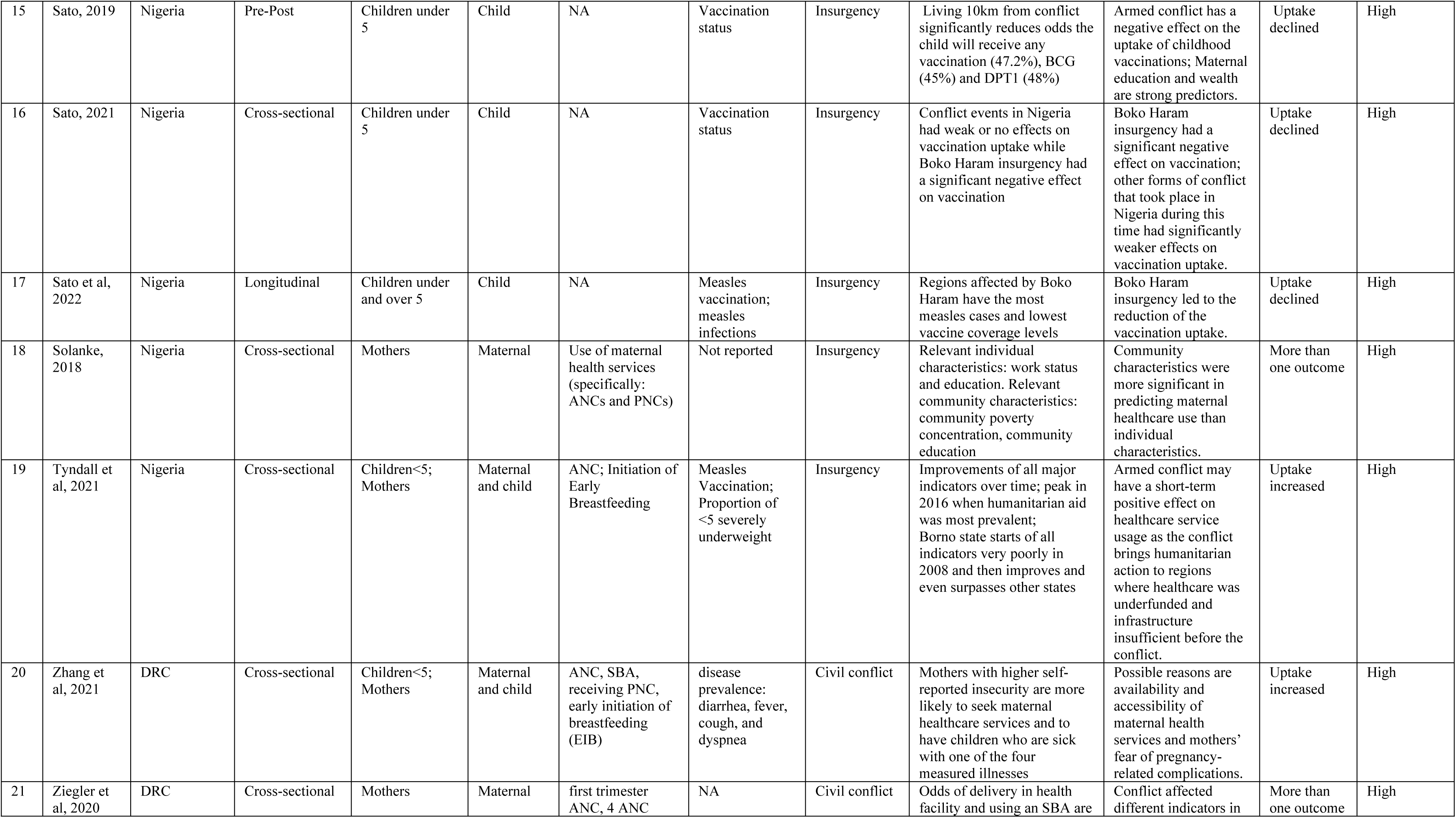
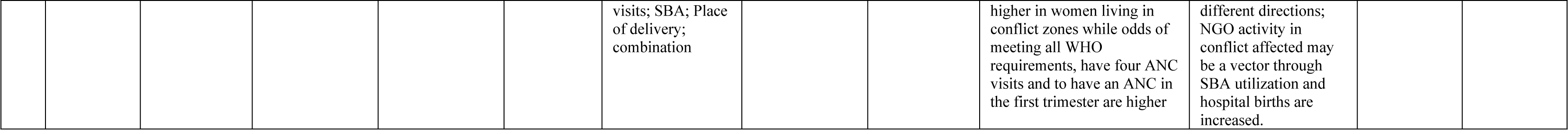
Summary of study findings.

| No | Author | Country | Study Design | Population | Types of HSB | Types of maternal health | Types of child health | Nature of armed conflicts | Result | Result Effects | Primary Outcomes | Quality of Evidence (GRADE) |
| --- | --- | --- | --- | --- | --- | --- | --- | --- | --- | --- | --- | --- |
| 1 | Ahmed et al, 2020 | Somalia | Cross-sectional | Women and children (Under 5) | Maternal and child | ANC, CPR & Caesarean section rate | DPT, measles and case management for pneumonia | Armed conflict | Increase of most intervention coverage in Somali land; ANC decrease in Puntland | Conflict decreased ANC attendance in Puntland but not in Somali land | More than one outcome | Moderate |
| 2 | Babakura et al, 2021 | Nigeria | Other (retrospective) | Children (Under 5) | Child | NA | Measles vaccine uptake | Insurgency | Measles vaccination of 68% of targeted children | Higher coverage in LGAs without security challenges | Intervention effective | High |
| 3 | Bayo et al, 2021 | South Sudan | Cross-sectional | Pregnant women and neonates | Maternal and child | ANC, health facility birth | Neonatal complications | Civil conflict | ANC visits declined by 21%; health facility births declined by 6.9% | Significant decline in key maternal and neonatal health service utilization indicators in Torit Hospital during conflict exposure | Uptake declined | High |
| 4 | Casey et al, 2013 | Uganda | Pre-Post | Women | Maternal | FP use and unmet need for FP | NA | Armed conflict | Increase in use of modern FP methods; increase in use of long-acting and permanent methods; decrease in % of women with unmet need for FP | Women will make use of FP services in conflict settings when provided | Uptake increased | High |
| 5 | Casey et al, 2017 | DRC | Pre-Post | Women | Maternal | FP | NA | Armed conflict | Increase in use of modern FP methods and long acting and permanent methods | Demand for FP methods, is present in humanitarian settings, and will be used when available | Uptake increased | High |
| 6 | Chi et al, 2015a | Burundi & Uganda | Other (Qualitative study) | Women of reproductive age | Maternal | SBA, ANC, CPR & Unmet need for FP | NA | Armed conflict | Factors affecting maternal health services utilization: financial barriers; level of household, community, and facility support; proximity; community perceptions | Conflict affects maternal health services utilization in diverse ways | Uptake remained unchanged | High |
| 7 | Chi et al, 2015b | Burundi & Uganda | Other (Qualitative study) | Women | Maternal | SBA, HIV/AIDS | NA | Armed conflict | Limited access to and poor quality of maternal and reproductive health services due to conflict. Abduction of health workers in Northern Uganda. Killing of health workers and ethnic bias in Burundi | Poor or no access to skilled services during the conflict, which has many perceived consequences | More than one outcome | High |
| 8 | Chukuma et al, 2019 | Nigeria | Cross-sectional | Women of reproductive | Maternal | ANC, health facility birth & | NA | Armed conflict | Reduction of ANC use by 13%, use of 4 ANC visits by | Boko Haram insurgency reduced the | Uptake declined | High |
|  |  |  |  | age |  | SBA |  |  | 35%, deliveries in health facilities by 38%, and skilled birth attendance by 26% during conflict. | probability of any antenatal care visits, of delivery at a health center, and of delivery by a skilled health professional. |  |  |
| 9 | Chukuma et al, 2021 | Kenya | Cross-sectional | Pregnant women | Maternal | ANC | NA | Civil conflict | When an initial ANC visit occurs in a facility within 10,000m of any conflict event, the client receives fewer recommended components of care. | The study demonstrated that conflict is an important determinant of non-utilization of ANC and the quality of care that women receive. | Other | High |
| 10 | Druetz et al, 2022 | Burkina Faso | Quasi-Experimental | Pregnant women | Maternal | Caesarian section rate, assisted delivery and ANC | NA | Insurgency | 7.7% reduction of caesarean sections; 2.5% reduction of assisted deliveries; 1.8% reduction of ANC visits. | Insecurity level in a community is negatively associated with the use of maternal healthcare services. | Uptake declined | High |
| 11 | McGinn et al, 2011 | Sudan; Uganda; DRC | Cross-sectional | Women of reproductive age | Maternal | Contraceptive use; knowledge of contraceptives | NA | Armed conflict | Low knowledge of contraceptive methods; Low use of modern contraceptive methods. | Low level of contraceptive use and knowledge associated with poverty, conflict, and low level of school attendance | Uptake declined | Moderate |
| 12 | Namasivayam et al, 2017 | Uganda | Cross-sectional | Women of reproductive age | Maternal | Contraceptive use; ANC; prenatal care; SBA; Institutional delivery | NA | Civil conflict | Lower odds for contraception use and institutional delivery for women in the north of Uganda across time; Higher odds of SBA in Northern Uganda. | Conflict generally has a negative effect on healthcare utilization though some indicators show the opposite development: higher odds of SBA in the north; this may be due to targeted humanitarian interventions in the crisis region. | Uptake increased | High |
| 13 | Nattabi et al, 2011 | Uganda | Cross-sectional | Patients; HIV Clinic Attendees | Maternal | Use of Family Planning Services | NA | Insurgency | 96% knew at least one FP method; 38% are currently using FP; low number of doctors in the region as a structural barrier to FP | Very low use of family planning methods despite high level of knowledge. | Other | High |
| 14 | Ojeleke et al, 2022 | Nigeria | Cross-sectional | Children <5; women of reproductive age | Maternal and child | Childbirth at health facilities and ANC visits | vaccination | Insurgency | Exposure to conflict events significantly lower odds of completing 4 ANC visits, birth in a health facility and receive child immunization | conflict has a negative impact on healthcare utilization. | Uptake declined | High |
| 15 | Sato, 2019 | Nigeria | Pre-Post | Children under 5 | Child | NA | Vaccination status | Insurgency | Living 10km from conflict significantly reduces odds the child will receive any vaccination (47.2%), BCG (45%) and DPT1 (48%) | Armed conflict has a negative effect on the uptake of childhood vaccinations; Maternal education and wealth are strong predictors. | Uptake declined | High |
| 16 | Sato, 2021 | Nigeria | Cross-sectional | Children under 5 | Child | NA | Vaccination status | Insurgency | Conflict events in Nigeria had weak or no effects on vaccination uptake while Boko Haram insurgency had a significant negative effect on vaccination | Boko Haram insurgency had a significant negative effect on vaccination; other forms of conflict that took place in Nigeria during this time had significantly weaker effects on vaccination uptake. | Uptake declined | High |
| 17 | Sato et al, 2022 | Nigeria | Longitudinal | Children under and over 5 | Child | NA | Measles vaccination; measles infections | Insurgency | Regions affected by Boko Haram have the most measles cases and lowest vaccine coverage levels | Boko Haram insurgency led to the reduction of the vaccination uptake. | Uptake declined | High |
| 18 | Solanke, 2018 | Nigeria | Cross-sectional | Mothers | Maternal | Use of maternal health services (specifically: ANC and PNCs) | Not reported | Insurgency | Relevant individual characteristics: work status and education. Relevant community characteristics: community poverty concentration, community education | Community characteristics were more significant in predicting maternal healthcare use than individual characteristics. | More than one outcome | High |
| 19 | Tyndall et al, 2021 | Nigeria | Cross-sectional | Children<5; Mothers | Maternal and child | ANC; Initiation of Early Breastfeeding | Measles Vaccination; Proportion of <5 severely underweight | Insurgency | Improvements of all major indicators over time; peak in 2016 when humanitarian aid was most prevalent; Borno state starts of all indicators very poorly in 2008 and then improves and even surpasses other states | Armed conflict may have a short-term positive effect on healthcare service usage as the conflict brings humanitarian action to regions where healthcare was underfunded and infrastructure insufficient before the conflict. | Uptake increased | High |
| 20 | Zhang et al, 2021 | DRC | Cross-sectional | Children<5; Mothers | Maternal and child | ANC, SBA, receiving PNC, early initiation of breastfeeding (EIB) | disease prevalence: diarrhea, fever, cough, and dyspnea | Civil conflict | Mothers with higher self-reported insecurity are more likely to seek maternal healthcare services and to have children who are sick with one of the four measured illnesses | Possible reasons are availability and accessibility of maternal health services and mothers' fear of pregnancy-related complications. | Uptake increased | High |
| 21 | Ziegler et al, 2020 | DRC | Cross-sectional | Mothers | Maternal | first trimester ANC, 4 ANC | NA | Civil conflict | Odds of delivery in health facility and using an SBA are | Conflict affected different indicators in | More than one outcome | High |
|  |  |  |  |  |  | visits; SBA; Place<br>of delivery;<br>combination |  |  | higher in women living in<br>conflict zones while odds of<br>meeting all WHO<br>requirements, have four ANC<br>visits and to have an ANC in<br>the first trimester are higher | different directions;<br>NGO activity in<br>conflict affected may<br>be a vector through<br>SBA utilization and<br>hospital births are<br>increased. |  |  |

The included studies reported a variety of outcomes for healthcare-seeking behaviors, with no overall consistent effect detectable. Ten studies highlighted a significant decline in the utilization of health services for both maternal and child health, while three reported an increase in the uptake of various healthcare services, and four others reported a mixed outlook (uptake for maternal health care services and decline in utilization in some areas). Factors influencing healthcare-seeking behavior in armed conflict settings included gender, marital status, the influx of humanitarian aid, nature and intensity of armed conflict, distance to conflict areas, as well as individual, community, and contextual factors. Table 5 provides a key synopsis and summary of the findings from all assessed articles.

In addition to the direct effect of armed violent conflict on maternal and child health behavior (i.e. the primary outcomes), it was observed that maternal and child healthcare utilization is affected by a variety of factors. Security challenges were found to affect service delivery and uptake [52,58,61]. Healthcare services and their quality were negatively affected by factors such as insufficient financing, shortage of healthcare personnel, and destruction of infrastructure [20,58]. Poor quality of healthcare services as well as past unpleasant experiences in health facilities, contributed to a lower demand for maternal healthcare usage [45,50,51]. Male partner’s attitudes or their negative perceptions of healthcare services emerged as significant barriers to positive health-seeking behavior [45,55,57,60]. Individual and community education, socio-economic status or well-being, and the act of watching TV were identified as predictors of health service uptake [60].

### Risk of bias and quality of evidence

The quality of studies in the systematic review was rated using GRADE, as recommended by the Cochrane Handbook for Systematic Reviews of Interventions [64,65]. This involved categorizing the quality of evidence into four levels: high, moderate, low, or very low. For each of the included studies, the evidence-level was determined, and the risk of bias assessed. Specifically, we ascertained that outcomes were a direct result of the investigation and that the process was scientific and reproducible. We also considered bias due to selection, detection, attrition, and reporting, and assessed quality with regard to imprecision, inconsistency, and indirectness [34]. The review was guided by an independently designed instrument based on GRADE guidance [36,66], reproduced in figure 3.

**Figure 3:**
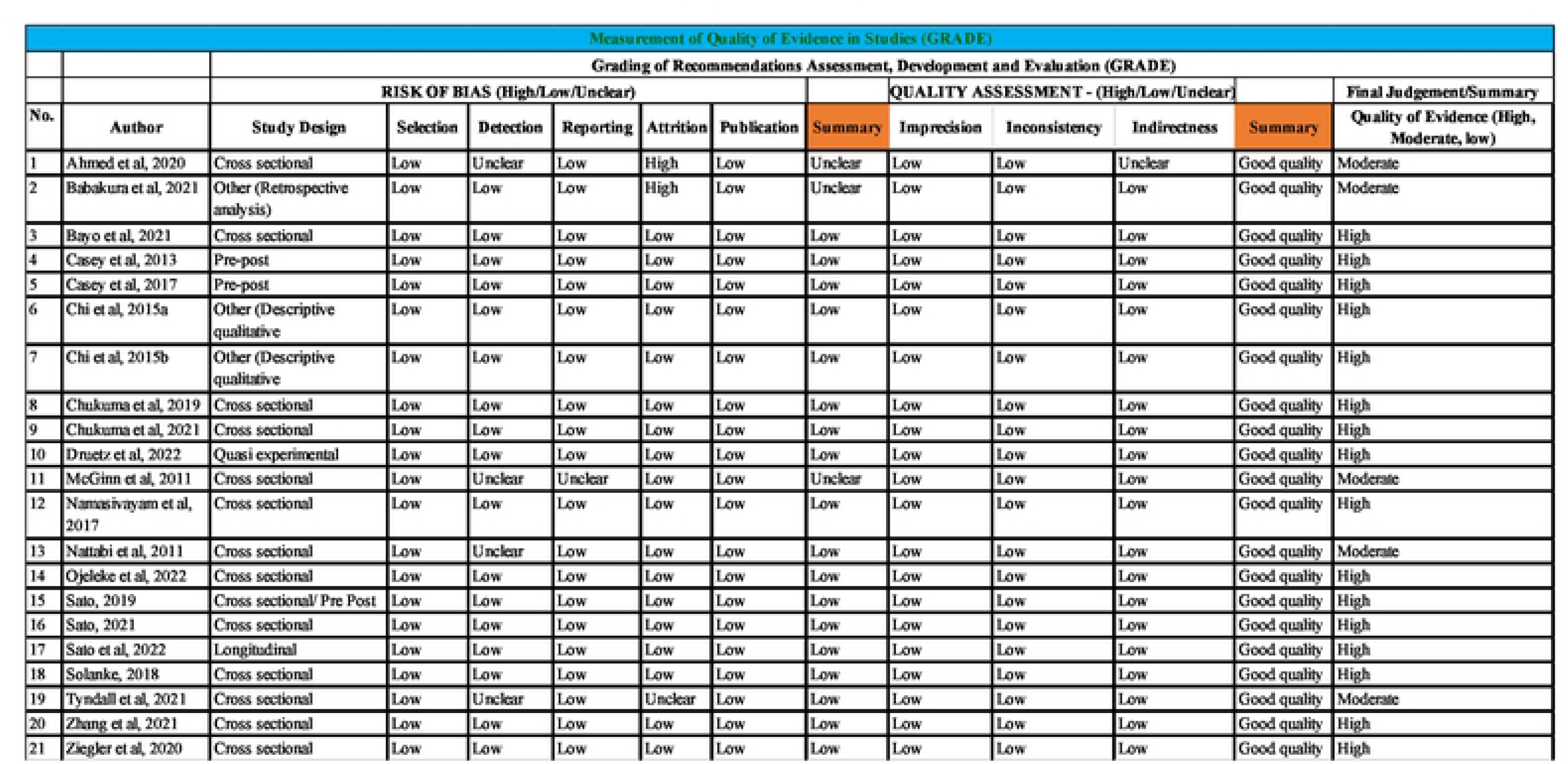
Quality of evidence and risks of bias in included studies.

The results of these assessments were summarized in the “Quality of evidence” column of the summary findings table. All the studies, with the exception of five, were considered to be of high quality. The studies deemed to be of moderate quality showed signs of detection, reporting biases, attrition, and indirectness in quality [39,53,55,58,61].

## DISCUSSION

A first notable observation is that all 21 included studies reported different outcomes for healthcare-seeking behaviors in SSA. In other words, there appears to be no shared standard as to what health outcomes to report in the context of armed violence. Second, while the majority of studies reported a significant *decline* in the utilization of health services for both maternal and child healthcare, a few reported an *increase* in the uptake of various healthcare services, with a minority reporting mixed outcomes. That is, we do not find a consistent effect of violent conflict on healthcare-seeking behavior. Rather, the studies reveal a more complex picture. However, factors such as marital status, male partner’s attitude, a woman’s education, income, media (act of watching television), education, humanitarian aid, nature and intensity of armed conflict and poverty level of communities were identified as determinants influencing health-seeking behavior and utilization in conflicts and post conflict settings in SSA. We first discuss findings related to healthcare-seeking behavior for maternal health services, and then transition to healthcare-seeking behaviors for child health services.

### Healthcare-seeking behavior for maternal health services

Three studies demonstrated a significant increase in healthcare-seeking behaviors, primarily related to the uptake of family planning, antenatal care, skilled attendance at birth, and postnatal care services [41, 47, 59]. For example, Casey et al. [47] reported a rise in the utilization of family planning commodities from 7.1% to 22.6% in northern Uganda and from 3.1% to 5.9% in the Democratic Republic of Congo. Similarly, in the Democratic Republic of Congo, mothers reporting higher insecurity were more inclined to seek maternal and child healthcare services, including professional ANC services, skilled attendants at delivery, postnatal care, early initiation of breastfeeding, and early care seeking for common childhood illnesses [59]. The increase in maternal health utilization in armed violent conflict situations might appear counterintuitive, but there are plausible reasons for its likelihood. The experience of war carnage or losses linked with armed conflicts could cultivate a greater appreciation for life (mother) and/or a heightened sense of protection for new life (pregnancy of infants), thereby promoting positive health-seeking behavior [1, 67, 68]. The human loss during armed conflict might also increase reproductive activity, be it out of necessity to compensate for the losses or for other reasons.

In contrast, approximately half (10) of the studies documented a significant decrease in healthcare-seeking behaviors for maternal health services. The most commonly affected health-seeking behaviors included the uptake of family planning, antenatal care, skilled attendance at birth, and health facility birth. Cesarean section rates and utilization of postnatal care services were also impacted. Antenatal care attendance showed declines in South Sudan, Nigeria, and Burkina Faso by 27.9%, 13%, and 1.8%, respectively [20, 50, 52]. Similarly, health facility births in South Sudan and Nigeria decreased by 6.9% and 38%, respectively, while skilled attendance at birth in Nigeria and Burkina Faso declined by 26% and 2.5%, respectively [20, 52]. A cross-sectional survey across three African countries affected by armed conflicts (Sudan, Uganda, and DRC) revealed a reduction in the use of family planning services by up to 16.2% [53]. Other studies reported an overall reduction in family planning, antenatal care, postnatal care, cesarean section rates, and the quality of antenatal care provided to clients [51–55, 57].

Interestingly, four studies reported mixed outcomes for maternal health service utilization even with the same study. For instance, ANC attendance increased from 30.2% to 51.2% in Somali land but declined from 30.1% to 27.9% in Puntland [39]. In northern Nigeria, the odds of undertaking four or more ANC visits are higher in conflict clusters than non-conflict clusters while exposure to conflict reduces the odds of a woman giving birth at a health facility [56]. Similarly, exposure to conflict events also lowered child immunization uptake [56]. Another study in the same region of northern Nigeria (Borno state) showed low levels of all maternal and child health indicators before the start of the insurgency. However, over time, all indicators improved [58]. In the DRC, women living in high or extremely high conflict zones were more likely to deliver in a health facility or have skilled attendance at birth, yet they were less likely to attend ANC than those living in moderate conflict zones [60]. This shows that the impacts of armed violent conflicts are context-specific, leading to underutilization of maternal health services in some areas and increased utilization in others. Also, different health seeking behaviors were observed for pre and during conflict periods. Therefore, interventions to address the health disparities must reflect the context of the conflicts and the respective periods.

### Healthcare-seeking behaviors for child health services

Similarly mixed outcomes were obtained with regard to healthcare-seeking behavior for child health services. Three studies reported that healthcare-seeking behavior for child health improved despite exposure to armed conflict events. The type of healthcare-seeking behaviors found to improve were vaccination (Diphtheria-Tetanus-pertussis (DPT), Measles and OPV), case management for pneumonia, and an overall increase in the use of healthcare services [39,59,61].

Conversely, five included studies revealed an overall decline in healthcare-seeking behaviors for child health in the region. The observed declines included vaccination rates (BCG, DPT, and Measles), the proportion of well-nourished under-fives, and care seeking for neonatal complications [43,56,58,62,63]. Specifically, in three states (Adamawa, Borno and Yobe) heavily affected by insurgency in northern Nigeria, it was found that if a child resides within 10km of a conflict zone, the odds were 47.2% lower for receiving vaccination overall, 45% lower for BCG, and 48% lower for DPT1 [43]. In the same states, the Boko Haram insurgency was found to have a substantial negative effect on vaccination, with the likelihood of a child ever getting vaccinated decreasing by 40% if born within the conflict areas [62]. Similarly, a geographical and time trend analysis of measles incidence and vaccination coverage in Borno and Yobe states (Nigeria) confirmed that exposure to violent activities by Boko Haram led to a reduction in vaccination uptake. The regions affected by insurgency exhibited the highest measles cases and lowest vaccine coverage levels [63].

Health seeking behavior often reflects the prevailing maternal and child health conditions, with these factors interacting in an interdependent manner to create a dynamic pattern of healthcare-seeking that remains flexible and susceptible to change [69]. The health system in sub-Saharan Africa is facing many challenges and exposure to armed conflicts is well-known to frequently disrupt of health service delivery, significantly affecting healthcare-seeking behaviors, especially among the most vulnerable groups – women and children. Therefore, research efforts should investigate how armed conflicts affect demand and supply of health services. Specifically, it is essential to explore how changes in healthcare-seeking behavior during armed conflicts contribute to the overall maternal and child morbidity and mortality rates in SSA [4].

### Factors influencing health-seeking behavior in armed conflict and post-conflict settings

During armed violent conflicts, maternal and child healthcare utilization is affected by a variety of factors, illustrated in Figure 4. First, at a structural level, exposure to armed conflict negatively impacts the delivery of healthcare services and service quality due to insufficient financing, lack of healthcare personnel, and infrastructure destruction [20,52,58,61].

**Figure 4:**
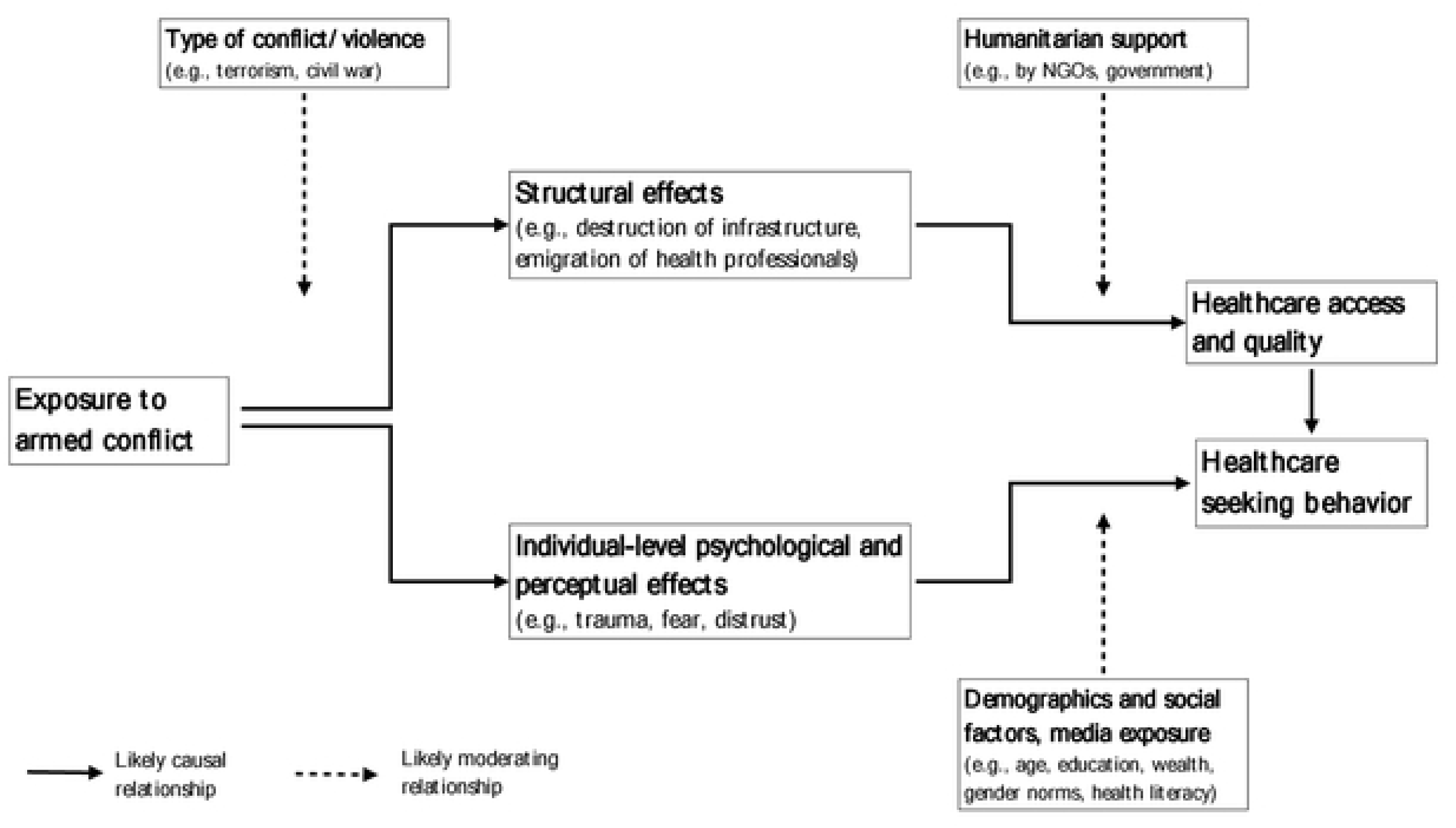
Factors linking exposure to armed conflict to health-seeking behavior.

Second, alleviating these overall negative impacts, humanitarian interventions and aid deliveries were found to be an important catalyst for healthcare-seeking behavior. Seven of the included reviewed papers highlighted the influx of humanitarian aid to armed conflict zones served as a possible vessel for improved maternal and child health utilization [39,45,54,56, 58, 59, 60]. Non-Governmental Organizations (NGOs), both international and local, provide health services, thereby improving health service delivery. Some conflict-affected areas did not have good healthcare services before the conflict, making the influx of humanitarian aid an improvement to maternal healthcare services, thereby making them both available and accessible [58,59]. The increase in healthcare services due to humanitarian intervention also had a positive effect on childhood vaccination. For instance, in Borno state (Nigeria), vaccination coverage rose from 12% in 2012 to 58% in 2016 [58]. It is important to note that these improvements resulting from humanitarian aid intervention are often not sustainable. When humanitarian aid departs after the end of a conflict, it is possible that the health state before the war may return, partially due to donor dependence and lack of a sustainability plan [39].

Apart from these factors influencing healthcare-seeking behavior by reducing access, a third relevant factor in maternal healthcare utilization that emerges is gender, as discussed in five of the examined papers [39,45,55,57,60]. The dominance of male decision-makers in healthcare choices, influenced by patriarchy or excessive masculinity, impacts family planning and household decisions. It also affects the uptake of ANC, PNC, or the use of SBA [39,45,55,57,60,70]. Even in emergency caesarian-sections, seeking permission from male heads of household can have fatal consequences for both the mother and her newborn. Male opposition serves as a primary barrier to modern family planning, stemming from prejudice, safety concerns, or the desire for a larger family, especially after losing family members to the conflict [39]. Male partner opposition is also a significant barrier to HIV counselling and testing [45,55].

Being married is negatively associated with taking up ANC and PNC visits and using a skilled birth attendant [57, 60]. This effect is lessened when the partner has higher education or when household healthcare-decision-making power is not male dominated. In patriarchal settings, women also tend to receive less favorable treatment, experience delays, or not receive care at all when not accompanied by their husband to appointments [45]. These situations are complicated during armed conflicts where husbands are deployed to defend the community or prosecute a war. Masculinity, as a significant determinant of healthcare-seeking behavior in SSA and an emerging area of global health research, have been strongly linked to child health outcomes [70–72]. However, this review shows that its effects extend beyond child health to include maternal health and possibly other health-seeking behavior within households in SSA. Further empirical research is required on this subject, especially focusing on maternal health, to guide appropriate behavior-change interventions.

A fourth set of factors influencing healthcare-seeking behavior in conflict settings is related to individual attitudes of those seeking health, such as heightened perceived health risks among women. Two of the studies showed that women in conflict areas were more likely to deliver in hospitals, use early term ANC services, and attend more ANC visits or use an SBA [54,60]. In other words, many expectant mothers in armed conflict zones seek healthcare services, and this effect appears to be due to perceived and heightened risks of unattended delivery [59,60]. Conversely, prior experiences of poor-quality healthcare services and unpleasant caregiver experiences (including the attitudes of healthcare workers) in health facilities in conflict-affected areas may negatively affect healthcare-seeking behavior [45,50,51].

Finally, in terms of demographic and community factors influencing healthcare-seeking behavior in areas exposed to armed conflict, women’s level of education and household income were positive predictors of healthcare service uptake, alongside media exposure (watching television), which served as a means of providing awareness and general health promotion [57,60]. At the community level, education had a positive, and poverty a negative impact on indicators of healthcare utilization [57].

### Implication of the experience of violent conflicts on maternal and child health outcomes

Armed violent conflict has mixed effects on maternal healthcare utilization in SSA. Several contextual factors contribute to the direction of these effects, including gender norms, service quality, safety, access to health facilities, and fear of complications during pregnancy, etc. There is a need for more studies to investigate these factors further. Similarly, armed conflict has been found to have a mostly negative but mixed effect on child health-seeking behavior in sub-Saharan Africa. Studies have shown that several indicators of child health and healthcare utilization worsened during the ongoing conflict [56]. However, some studies show an upward trend as well, e.g., increased healthcare services due to humanitarian aid in conflict regions, specifically in regions where coverage was not extensive before the conflict [61]. Overall, it can be concluded that armed conflict negatively impacts child health. However, these findings suggest that with targeted health provision campaigns, children in conflict-affected areas can be reached, and their access can even be improved.

### Relationship between armed violent conflict and vaccination uptake

Out of the six papers discussing childhood immunization, four concluded that armed conflict decreases vaccination uptake, one concluded that it increases vaccine uptake through increased availability owing to factors identified previously (humanitarian intervention), and one did not discuss the relationship. The general detrimental effect of armed violent conflict on childhood immunization is clear; nevertheless, this relationship is influenced by several factors. Security concerns and inaccessibility are major factors [61]. The Boko Haram insurgency decreased the odds of childhood vaccination by 35%, moderated by maternal education and household income [43,62]. Armed violent conflicts rarely affected childhood vaccine uptake when the mother had some educational attainment, but it decreased the odds of vaccination by 64.3% if the mother was uneducated [62]. Importantly, income moderated the relationship in opposing ways. While armed conflicts did not affect the odds of vaccination for children from a poor household, those growing up in a non-poor household were 41.8% less likely to get vaccinated when exposed to armed violent conflict. This might be due to the prevalence of Boko Haram insurgency in wealthier or urban areas compared to rural ones.

### General implication of armed violent conflict for population health

Armed conflict in SSA significantly impacts maternal and child healthcare-seeking behaviors. The study has shown that armed conflict leads to reduced health service coverage and utilization, affecting reproductive, maternal, newborn, and child health interventions, nutritional status, and even child mortality. The escalation of armed conflict in the region undermines public infrastructure, including health systems, making it challenging to maintain adequate healthcare services. However, we have also seen that armed conflict may unintentionally lead to positive effects on maternal health-seeking behaviors in regions with poor health infrastructure, especially when humanitarian health assistance is provided [54,56,58–60]. This highlights the complex and multifaceted influence of armed conflict on healthcare-seeking behaviors for maternal and child health in sub-Saharan Africa, emphasizing the need for targeted policy interventions and humanitarian assistance to mitigate the negative impact of armed conflict on healthcare-seeking behaviors in the region.

There is also a clear need for further research to fill remaining gaps in our knowledge. A first area where research is needed is the contextual determinants shaping the relationship between exposure to armed conflict and healthcare-seeking behavior. While humanitarian aid seems to be one factor moderating the effect of armed conflict on healthcare-seeking behavior, it is unlikely to explain the full variation in findings. Another possible factor is the type of violence vulnerable populations are exposed to. Prior research has established that terrorism has strong negative effects on child health [25,43], and it is noteworthy that some of the strongest reductions in healthcare-seeking behavior were in areas affected by attacks by Boko Haram, a known terrorist organization that explicitly and repeatedly has targeted civilians. It is possible that other forms of violence, such as insurgencies fought mainly between rebels and government forces, have less adverse effects [43]. These and other potential explanations for the mixed outcomes described in this review will need to be tested in further studies.

Another aspect that has so far received little attention is the effects of exposure to violent conflict on individual-level perceptions and psychological dispositions, including trauma, and their potential knock-on effects on healthcare-seeking behavior. Only six studies discuss subjective factors at all [19,45,50,53,55,59], and none explicitly deals with perceptions of fear and psychological trauma. This is despite the fact that aspects such as increased fear of future violence and lost trust in state institutions have been hypothesized to drive many of the detrimental changes in healthcare-seeking behavior [25]. Explicitly exploring these channels remains an important area for future research.

As with any review, this study has some limitations. The search strategy might have missed relevant studies due to the broad concept of healthcare-seeking behaviors for maternal and child health. Additionally, including only published studies in English from SSA limits our ability to provide a global narrative on the influence of armed conflict on healthcare-seeking behaviors for maternal and child health. Moreover, the absence of any randomized controlled trial (RCT) in our review is noteworthy, considering that RCTs are considered the gold standard in healthcare research. This absence once again underlines the need for further, high-quality research in this area to better inform evidence-based decision-making.

## CONCLUSIONS

This review sheds light on the intricate and varied impact of armed conflict on healthcare-seeking behaviors for maternal and child health in SSA, highlighting the need for targeted policy interventions and increased humanitarian assistance to mitigate negative effects. It serves as a valuable academic and policy resource for understanding population health behaviors within conflict and post-conflict contexts in SSA. Additionally, it represents an initial attempt to grasp and contextualize the influence of exposure to armed conflict on healthcare-seeking behaviors for both maternal and child health services in the region. While previous reviews have primarily focused on the effects of armed conflicts on infrastructure, healthcare systems, and service coverage, the impact of armed conflicts on behavioral adjustments among healthcare-seekers has received less attention. This is concerning, as prompt healthcare-seeking is critical for managing existing health problems effectively. Therefore, understanding the factors shaping healthcare-seeking behaviors in the context of armed violence is crucial for preventing harm to vulnerable populations, including children and mothers.

Alongside providing suggestions for further research, this review will help policymakers and development practitioners or researchers narrow the scientific knowledge gap on how exposure to armed violent conflict or post-conflict situations interacts with healthcare-seeking behaviors for maternal and child health in SSA. It also contributes to a deeper exploration and description of micro-level factors shaping healthcare-seeking behaviors in armed conflict settings within the sub-region, through the lenses of available evidence. Armed violent conflict has mixed effects on child and maternal healthcare utilization in the region, indicating a need for further studies to elucidate the interrelated factors. While armed conflict disproportionately impacts child health compared to maternal health, it is noteworthy that, exposure to armed violent conflict may unintentionally lead to positive effects in regions with poor health infrastructure, namely if humanitarian health assistance is unhindered.

## LIST OF ABBREVATION

AJOL: African Journals Online
ANC: Antenatal Care
BCG: Bacillus Calmette–Guérin vaccine
BH: Boko Haram
CRD: Centre for Reviews and Dissemination of Systematic Reviews
CPR: Contraceptive prevalence rate
DPT: Diphtheria-Pertussis-Tetanus vaccine
DRC: Democratic Republic of Congo
EIB: Early initiation of breastfeeding
FP: Family Planning
GRADE: Grades of Recommendation, Assessment, Development and Evaluation
ICRC: The International Committee of the Red Cross
IDP: Internally Displaced Person
LGA: Local Government Area
MCH: Maternal and Child Health
MeSH: Medical Subject Headings
NGO: Non-Governmental Health Organization
OPV: Oral polio vaccine
PICO: Population, Intervention, Comparison and Outcomes
PNC: Postnatal Care
PRISMA-P: The Preferred Reporting Items for Systematic Reviews & Meta-Analyses Protocols
SBA: Skilled birth attendant
SSA: Sub-Saharan Africa
UCDP: The Uppsala Conflict Data Program
UNDP: United Nations development Programme
WHO: World Health Organization

## DECLARATIONS

### Ethics approval and consent to participate

Ethical review and consent to participate are not applicable for this study, as this is a systematic review of previously published studies.

### Data availability statement

The datasets used and/or analyzed during the current study are available on https://osf.io/gnkyc/

### Author’s Contribution

Conceptualization: GCA, LA, and MS.

Methodology: GCA, LA, and MS.

Investigation: PMES and RIJ.

Writing – original draft: GCA, LA, PMES, RIJ, and MS.

Writing – review & editing: GCA, LA, and MS.

All authors have read and agreed to the published version of the manuscript.

## Data Availability

All relevant data are within the manuscript. In addition, other datasets used and/or analyzed are available on https://osf.io/gnkyc/

